# Combining genome-wide polygenic scores with registry data for colorectal cancer risk-based screening

**DOI:** 10.64898/2025.12.04.25341629

**Authors:** Anne Krogh Nøhr, Marie Giehm Overby, Mads Munk Nielsen, Rikke Hedegaard Jensen, Laurids Østergaard Poulsen, Ole Thorlacius-Ussing, Simon Ladefoged Rasmussen, Berit Andersen, Ismail Gögenur, Erik Sørensen, Ole Birger Vesterager Pedersen, Christian Erikstrup, Nanna Brøns, Michael Schwinn, Christina Mikkelsen, Mie Topholm Bruun, Malene Møller Jørgensen, Claus Anders Bertelsen, Jens Georg Hillingsø, Estrid Høgdall, Sisse Rye Ostrowski, DBDS Genomic Consortium, DCB Research Consortium, Martin Bøgsted, Rasmus Froberg Brøndum

**Affiliations:** Center for Clinical Data Science, Aalborg University and Aalborg University Hospital, Aalborg, Denmark; Clinical Cancer Research Center, Aalborg University Hospital, Aalborg, Denmark; Department of Clinical Medicine, Aalborg University, Aalborg, Denmark; Department of Oncology, Aalborg University Hospital, Aalborg, Denmark; Department of Gastrointestinal Surgery, Aalborg University Hospital, Aalborg, Denmark; Department of Public Health Programmes and University Clinic for Cancer Screening (UNICCA), Randers Regional Hospital, Randers, Denmark; Department of Clinical Medicine, Aarhus University, Aarhus, Denmark; Department of Clinical Medicine, Faculty of Health and Medical Science, University of Copenhagen, Denmark; Center for Surgical Science, Zealand University Hospital, Køge, Denmark; Department of Clinical Immunology, Rigshospitalet, Copenhagen University Hospital, Copenhagen, Denmark; Department of Clinical Immunology, Zealand University Hospital, Køge, Denmark; Department of Clinical Immunology, Aarhus University Hospital, Aarhus, Denmark; The Novo Nordisk Foundation Center for Basic Metabolic Research, Faculty of Health and Medical Sciences, University of Copenhagen, Copenhagen, Denmark; Department of Clinical Immunology, Odense University Hospital, Odense, Denmark; Department of Clinical Immunology, Aalborg University Hospital, Aalborg, Denmark; Department of Surgery, Copenhagen University Hospital – North Zealand, Hillerød, Denmark; Department of Digestive Diseases, Transplantation and General Surgery, Copenhagen University Hospital, Copenhagen, Denmark; Department of Pathology, Herlev Hospital, University of Copenhagen, Copenhagen, Denmark

## Abstract

**Background:** Aggregating effects of common genetic variants into a polygenic risk score (PRS) has shown potential for risk-based colorectal cancer (CRC) screening. However, its utility must be evaluated in ancestrally diverse populations with diverse tumor characteristics and compared to the current screening standard, the fecal immunochemical test (FIT).

**Objective:** Evaluate the utility of PRS for risk-based CRC screening.

**Design:** The cohort included 112,204 individuals from the Copenhagen Hospital Biobank (8,995 with adenoma and 9,246 with CRC), all with linked genetic and health registry data. A subset (N=20,658) also had available FIT results. CRC PRSs were evaluated for their association with lifetime adenoma and CRC risk and their predictive value individually and combined with FIT.

**Results:** PRS stratified population-calibrated lifetime adenoma and CRC risk independently of ancestry and sex. Incidence curves showed that individuals with a high PRS reached the incidence levels of low-PRS individuals around 10 years earlier, between ages 45 to 60. PRS also stratified risk across tumor location and histology but showed no association among individuals with deficient mismatch repair tumors (N=623). Predictive modeling indicated that combing PRS with FIT did not meaningfully improve prediction of adenoma or CRC at first screening, negative colonoscopy outcomes among FIT-positive participants, or adenoma or CRC occurrence within two years after a negative FIT.

**Conclusion:** PRS effectively stratifies lifetime adenoma and CRC risk and shows promise for guiding risk-based screening initiation and intensity. When combined with FIT, it adds limited additional predictive value.

## Introduction

Population-based colorectal cancer (CRC) screening programs have significantly reduced CRC incidence and mortality by enabling early detection and removal of precancerous adenomas^1–3^. These programs are typically offered biennially to adults aged 50 to 74, starting with a stool-based test, often the fecal immunochemical test (FIT), with positive results leading to confirmatory colonoscopy^2,4^. While colonoscopy is effective, it is invasive and costly, and stool-based tests like FIT have limited predictive performance. For instance, in Denmark, FIT misses ∼34% of CRC cases detected between screenings^5^, and only 50% of those with a positive FIT are diagnosed with adenomas or CRC, resulting in unnecessary colonoscopies^6^. Given these limitations, risk-based screening represents a promising alternative, with the potential to reduce unnecessary colonoscopies while improving detection rates.

Risk-based screening is already recommended for individuals with a family history of CRC, familial adenomatous polyposis (FAP), or hereditary non-polyposis colorectal cancer (HNPCC).^7^ FAP and HNPCC are caused by pathogenic variants with high penetrance^8^. Another component of CRC heritability lies in the common genetic variants with smaller effects, called single nucleotide polymorphisms (SNPs), identified by genome-wide association studies, which have already uncovered hundreds of CRC-associated variants^9–12^. Studies have demonstrated, that combining the effect of these variants into a polygenic risk score (PRS) can identify individuals of higher risk of CRC^13–22^. An appealing feature of PRS is that, since our genetics remain static, they can provide insights into lifetime risk. However, despite evidence supporting implementing PRS in risk-based screening, the approach has not been implemented.

To move towards clinical implementation several issues must be addressed. First, realistic lifetime risk estimates of colorectal adenomas and CRC requires recalibration to the general population, which has so far only been done in one study limited to individuals of European ancestry^23^. For PRS to be useful in a real-world screening, it must be generalizable across diverse ancestry groups. This is challenging, as the best-performing PRS methods require genetic data from all ancestries, which are not yet available. Consequently, most studies focus on Europeans, only few have evaluated PRS performance in minority ancestry groups^21,16^, and none have included individuals irrespective of ancestry.

Second, it remains unclear whether PRS can stratify risk among all CRC patients independent of different clinical or tumor characteristics. Only few studies have investigated this, for example, by age of onset^16,17,21,23,24^, sex^21,23,24^, tumor site^17,23^, histology type^23^, and presence of adenomas^23^. To our knowledge, no studies have examined the association between PRS and mismatch repair (MMR) status. This is relevant because while most CRCs are MMR-proficient (pMMR), a subset arise from MMR deficiency (dMMR), either sporadically or as part of HNPCC, and PRS may perform differently in these tumors, as one study found it not to be predictive of HNPCC^25^.

Finally, the clinical value of PRS should be assessed relative to the current FIT-based screening strategy. However, only a few studies have compared the two directly: one study found no improvement in diagnostic accuracy for detecting advanced colorectal neoplasia compared with FIT alone^26^, whereas another study, focusing on FIT-positive individuals, reported that PRS improved predictive performance^27^.

To address these gaps, we conducted a cohort study using data from 112,204 individuals in the Copenhagen Hospital Biobank (CHB)^28^ to address three objectives. First, we assessed the utility of PRS for stratifying lifetime risk of CRC and adenomas across sex and ancestry groups. Second, we examined whether PRS can provide clinically meaningful risk stratification among CRC patients, independent of tumor related characteristics. Finally, we evaluated the predictive performance of PRS in comparison with the currently implemented FIT-based screening strategy.

## Methods

### Patients

The study includes individuals from CHB, a large hospital cohort consisting of leftover ethylenediaminetetraacetic acid (EDTA) whole blood samples collected for blood typing and screening from outpatients or patients admitted to hospitals in the Capital Region of Denmark^28^. Each individual was linked to Danish national registries through their civil registration number. Registries include the Danish Cancer Registry (CAR)^29^, the Danish National Patient Register (DNPR)^30^, and the Danish Civil Registration System (CRS)^31^. The data was also linked to the Danish Colorectal Cancer Group Database (DCCG)^32,33^, established in 2001, which includes variables such as tumor type, histology, lifestyle factors, and comorbidities. Individuals were excluded if, at the time of data extraction, they were registered in the CRS as not residing in Denmark, missing, or if their civil registration number had been annulled, deleted, or changed. They were also excluded if they had withdrawn consent or if their genetic material failed quality control.

A subset of individuals was from 1 March 2014 invited to participate in the Danish screening program. Information on their participation in the screening program, FIT results, colonoscopy results etc., was obtained from the Danish Colorectal Cancer Screening Database (DCCSD)^34^. The same exclusion criteria apply to this subset. Furthermore, individuals were excluded if they died within one month of receiving an invitation or if they had a prior CRC diagnosis. A flowchart visualizing the available data is provided in Figure 1.

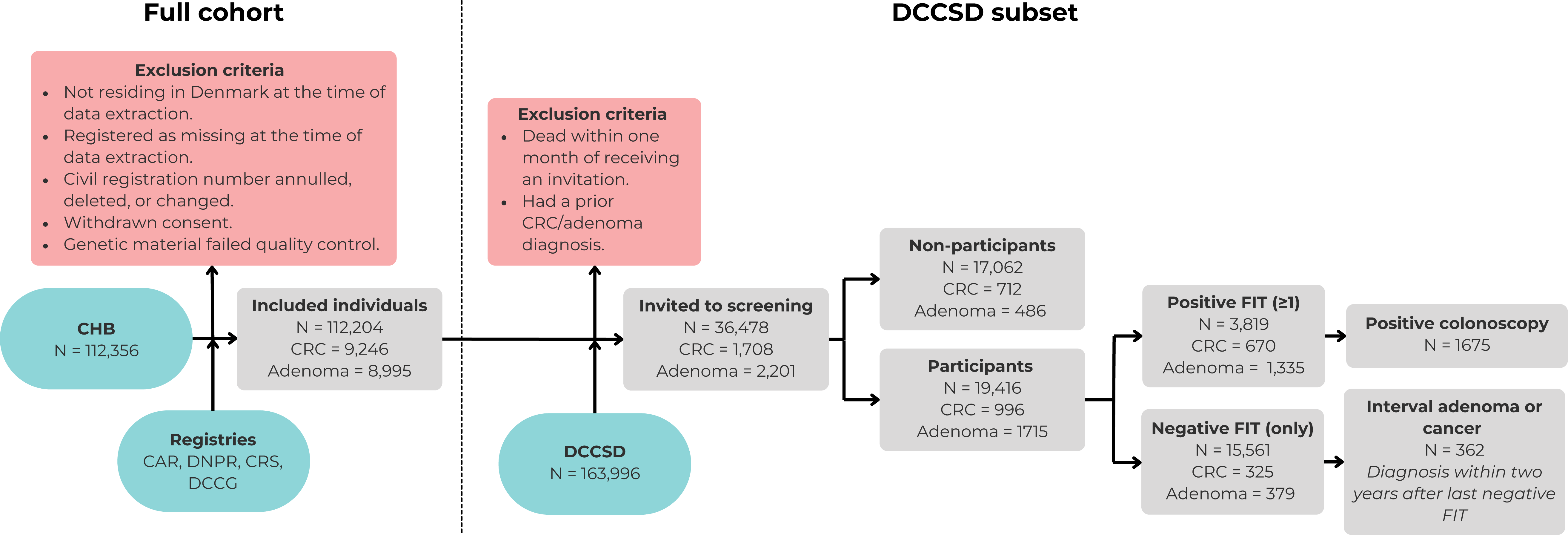

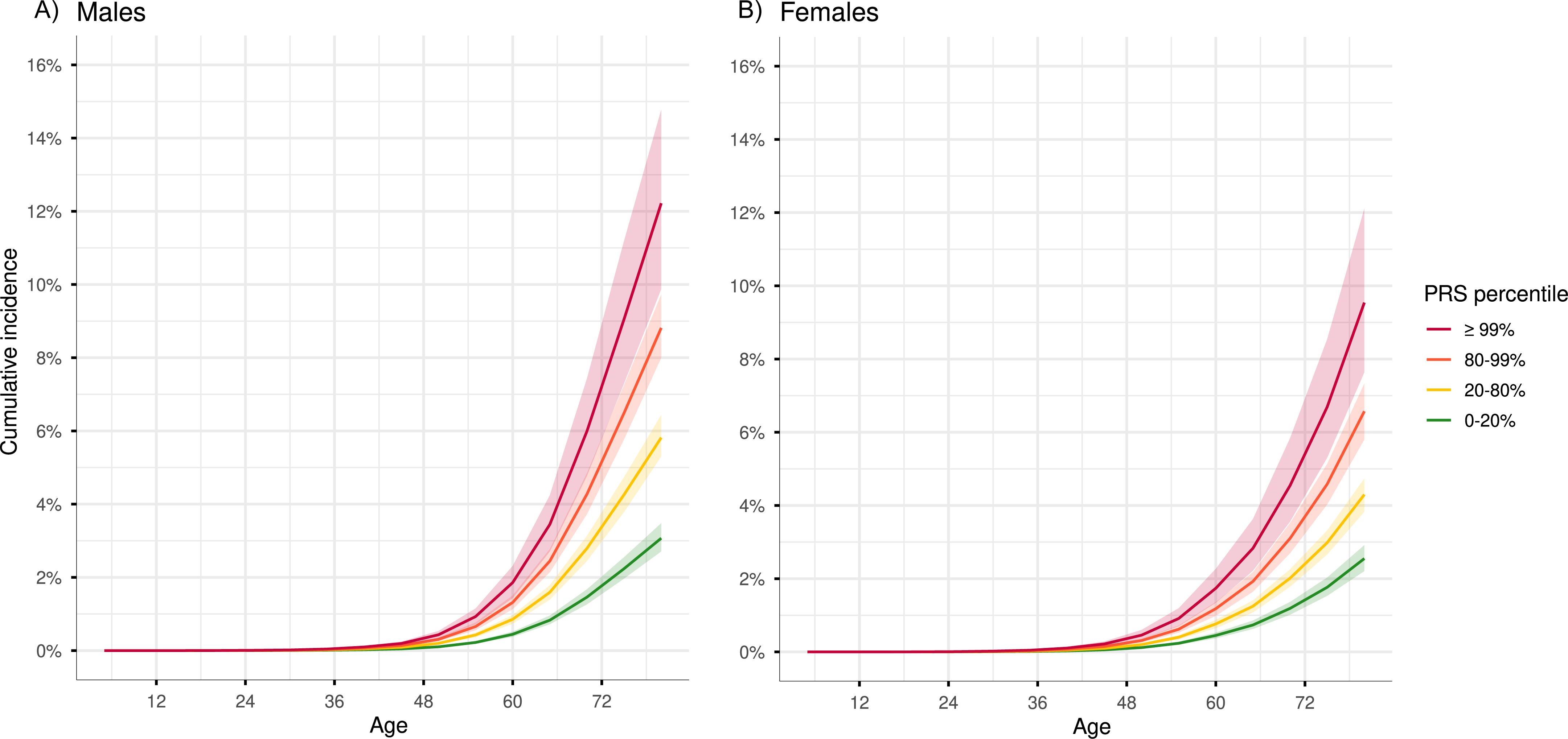

### Ethical approval

The study was approved by the Danish National Committee on Health Research Ethics in 2022 (NVK-2200966). Data processing has been registered with the Capital Region in accordance with Article 30 of the GDPR (P-2022-393). Patients or the public were not involved in the design, conduct, reporting, or dissemination of our research.

### Outcome and tumor and clinical characteristics

The primary outcome was the first CRC diagnosis. Individuals were defined as CRC cases if they had any CRC cancer diagnosis (ICD-10 codes C18-20, C26) in CAR or in DCCG. As a secondary outcome, colorectal adenomas were assessed and defined as the presence of an ICD-10 diagnosis code D12 (D12, D120-D126, D128, D129) in DNPR, also including hyperplastic polyps. Finally, to address the study’s last objective, we defined a combined outcome of CRC or adenoma, as it focuses on the FIT-based screening strategy, which aims to identify both adenomas and CRCs.

CRC tumors were described according to anatomical site, as recorded in the DCCG, and categorized as proximal colon, distal colon, rectum, or unspecified colon. The proximal colon included the caecum, ascending colon, hepatic flexure, and transverse colon, while the distal colon comprised the splenic flexure, descending colon, and sigmoid colon. MMR status (pMMR vs. dMMR) was also obtained from DCCG, where it is determined by immunohistochemical tests of biopsy material or surgical specimens to assess protein expression of MLH1, PMS2, MSH2, and MSH6. dMMR was defined as complete loss or markedly reduced expression in one or more MMR proteins in all tumor cells, with preserved expression in adjacent stromal cells and lymphocytes.

Positive and negative FIT results were defined as in the Danish Bowel Cancer Screening Program, using the threshold of 20 µg Hb/g feces ^5^, with a positive FIT being any value above this threshold. A positive colonoscopy is defined as at least one screening-derived colonoscopy that detects CRC or adenoma (also including polyps).

### Polygenic risk scores

PRS were estimated from imputed genotype data, obtained as described in Supplementary Material 1. In short, the EDTA whole blood samples were genotyped by deCODE Genetics using Illumina’s Infinium Global Screening Array v1 or v3. The genotype data underwent standard quality control and was imputed using deCODE genetics in-house imputation workflow^35^. Details on the estimation of principal components and genetic ancestry for each individual are provided in Supplementary Material 1.

For each individual, a PRS for CRC was calculated using weights from 203 of the 205 genome-wide significant SNPs identified in the largest CRC genome-wide association study to date^12^, applying PLINK2 (two variants were not available in our dataset). The PRS was constructed using only these top-associated SNPs, since genome-wide PRS approaches would require a reference linkage disequilibrium panel for all ancestries, a resource that is not yet available. Finally, the PRSs were standardized to have zero mean and unit variance.

Individuals were categorized into PRS groups: 0-20%, 20-80% (reference), 80-99%, and ≥99%, as per Tamlander et al.^23^ This approach was chosen due to the linear association between PRS and lifetime CRC risk, with no natural cutoffs for groupings (Supplementary Material 2, Supplementary Fig. 1).

### Statistical analysis

In this cohort study cumulative incidences and Cox proportional hazards models were used to evaluate lifetime risk of CRC and adenomas, with age as the time scale. In all analyses, left truncation was used to avoid immortal time bias. Thus, follow-up began at the outcome-specific entry date (listed in Supplementary Table 1) or at birth for individuals born after that date. For cases, follow-up ended at age of first CRC diagnosis (in the CRC analysis) or age of first adenoma (in the adenoma analysis). For the rest, follow-up ended at age of death, age at last record in the registries (2022-09-19) or age 80. An upper age limit of 80 years was chosen to account for competing mortality risks and changes with biological aging, ensuring more accurate and meaningful risk estimates. Furthermore, the models were adjusted for birth year, chip version, and ancestry by including the first four principal components. If the model was not stratified on sex, this was also included as a covariate.

#### Lifetime risk of CRC and adenomas across sex

For the PRS groupings, the lifetime risk of CRC was evaluated using cumulative incidence of CRC from birth to age 80, accounting for the competing risk of death from causes other than CRC. Since CHB is a hospital cohort and is thus not representative of the general Danish population the cumulative incidences were recalibrated for each sex using the method described by Jermy et al. 2024^24^ and using incidence, prevalence, and mortality estimates for Denmark from Global Burden of Disease (GBD) 2021 data^36^.

Lifetime risk of CRC and adenomas was also evaluated using Cox models to estimate HR for the PRS groupings. Cox models were fitted for each of the two outcomes: 1) using all individuals with and without an interaction term between sex and PRS, 2) stratified on Ancestry (European vs. non-European) and sex.

#### Association of PRS with lifetime CRC risk across clinical and tumor characteristics

Cause-specific Cox models were used to evaluate whether PRS can risk-stratify all CRC patients, independently of tumor characteristics. These characteristics included: CRC site, tumor MMR classification, and tumor histology. All these variables were categorical, each with at least two levels (e.g., the histology of CRC included the levels "adenocarcinoma," "mucinous adenocarcinoma", "poorly differentiated adenocarcinoma", and "other carcinoma"). A separate model was fitted for each specific level of the categorical variables, with the endpoint defined as CRC characterized by that level (e.g., CRC with the histology "adenocarcinoma"). The remaining level(s) were treated as competing risks and censored. Individuals with missing values for these characteristics were treated as controls.

### Predictive modelling

A subset of individuals was linked to DCCSD, and three analyses were conducted to assess whether the addition of PRS to FIT, compared with FIT alone, enhanced the performance of:

1. Prediction of CRC or adenoma risk from the time of an individual’s first screening participation.
2. Prediction of colonoscopy outcomes (positive vs. negative findings) among participants with a positive FIT.
3. Prediction of CRC or adenoma within two years following a negative FIT result.

In all three analyses model performance was evaluated for four distinct predictor sets: (1) a baseline set including age, sex, PC1-PC4, and genotyping chip; (2) baseline plus PRS; (3) baseline plus FIT value; and (4) baseline combined with both PRS and FIT. In all analyses, FIT was included as a continuous variable. Cox models were used in Analysis 1, and logistic regression was used in Analyses 2 and 3. In Analysis 3, to ensure that we did not simply use the first FIT for the controls but instead selected the most appropriate FIT to better match those of the cases, each case was matched with three controls based on sex and age at the time of the FIT. Model performance was evaluated using 10-fold cross-validation with an even proportion of cases in each split. The performance metric for Analysis 1 was the concordance index (C-index), whereas Analyses 2 and 3 were evaluated using the area under the receiver operating characteristic curve (AUC). A more detailed description is provided in Supplementary Material 3.

### Software

All statistical analyses were conducted using R version 4.4.0. Cox models were fitted with the coxph function from the survival package^37^. Cumulative incidence was estimated using the cuminc function from the cmprsk package^38^. Spline-based models were constructed using the rcs function from the rms package^39^.

## Results

### Cohort characteristics

A total of 112,204 individuals (10,039 with non-European genetic ancestry) from CHB were included in the study, of whom 9,246 were diagnosed with CRC (2,849 with rectal cancer and 6,397 with colon cancer). Furthermore, 8,995 individuals were diagnosed with a minimum of one colorectal adenoma. The median age of first CRC was 72 years (lower quartile (LQ): 64 years; upper quartile (UQ): 79 years) and 44% were female. The median age of first adenoma was 70 years (LQ: 63 years; UQ: 75 years) and 40 % were female. Demographics, clinical, and tumor characteristics of all individuals are provided in Table 1. The median follow-up time was 76.5 years (95%CI:76.4, 76.6) for CRC as the outcome and 76.4 years (95%CI:76.3, 76.5) for adenomas.

**Table 1.** Demographics, clinical, and tumor characteristics.

| **Characteristic** | **Colon (C18,26)**<br>**N = 6,397<sup>1</sup>** | **Rectum (C19-20)**<br>**N = 2,849<sup>1</sup>** | **Control**<br>**N = 102,958<sup>1</sup>** |
| --- | --- | --- | --- |
| **Gender** |  |  |  |
| Female | 3,073 (48%) | 1,025 (36%) | 51,307 (50%) |
| Male | 3,324 (52%) | 1,824 (64%) | 51,651 (50%) |
| **Birth year** | 1939 (1932, 1947) | 1941 (1933, 1949) | 1942 (1932, 1952) |
| **Year of diagnosis** | 2014 (2010, 2016) | 2013 (2008, 2015) |  |
| **CRC site** |  |  |  |
| Proximal colon | 2,356 (44%) | 0 (0%) |  |
| Distal colon | 2,307 (43%) | 0 (0%) |  |
| Unspecified colon | 717 (13%) | 0 (0%) |  |
| Rectum | 0 (0%) | 2,412 (100%) |  |
| Unknown | 1,017 | 437 |  |
| **ECOG<sup>2</sup> Performance Status** |  |  |  |
| 0 | 1,286 (45%) | 584 (53%) |  |
| 1 | 883 (31%) | 298 (27%) |  |
| 2 | 477 (17%) | 137 (13%) |  |
| 3 | 180 (6.3%) | 63 (5.8%) |  |
| 4 | 37 (1.3%) | 10 (0.9%) |  |
| Unknown | 3,534 | 1,757 |  |
| **Histology** |  |  |  |
| Adenocarcinoma | 2,611 (77%) | 1,301 (90%) |  |
| Carcinoma (Medullary, Signet ring cell, Undifferentiated) | 93 (2.7%) | 14 (1%) |  |
| Poorly differentiated adenocarcinoma | 317 (9.4%) | 56 (3.9%) |  |
| Mucinous adenocarcinoma | 364 (11%) | 81 (5.6%) |  |
| Unknown | 3,012 | 1,397 |  |
| **DNA mismatch repair (MMR)** |  |  |  |
| Deficient (dMMR) | 603 (19%) | 20 (1.5%) |  |
| Proficient (pMMR) | 2,585 (81%) | 1,309 (98%) |  |
| Unknown | 3,209 | 1,521 |  |
| **Adenoma diagnosis** |  |  |  |
| Adenoma | 1,240 (19%) | 485 (17%) | 7,270 (7%) |
| No adenoma | 5,157 (81%) | 2,364 (83%) | 95,688 (93%) |
| **Ancestry** |  |  |  |
| European | 5,903 (92%) | 2,610 (92%) | 93,652 (91%) |
| Non-European | 494 (7.7%) | 239 (8%) | 9,306 (9%) |
| **PRS strata** |  |  |  |
| 0-20 | 749 (12%) | 269 (10%) | 21,423 (21%) |
| 20-80 | 3,767 (59%) | 1,679 (59%) | 61,876 (60%) |
| 80-99 | 1,759 (27%) | 835 (29%) | 18,724 (18%) |
| ≥99 | 122 (2%) | 66 (2%) | 935 (1%) |

[1] n (%); Median (Q1, Q3), [2] ECOG = Eastern Cooperative Oncology Group.

### Lifetime risk of CRC in PRS groups and across sex

Population-recalibrated cumulative incidence showed that PRS identifies individuals at both higher and lower risk, and that incidence was higher in men than in women (Figure 1.A and 1.B). For men the cumulative incidence for CRC at age 80 was 12.22% (95%CI:9.87, 14.76) for individuals in the ≥99th and 8.81% (95%CI:8.00, 9.73) in the 80th-99th percentile group. In contrast, among individuals in the reference group (20th-80th percentile), 5.82% (95%CI:5.30, 6.44) developed CRC by age 80. For those in the 0-20th percentile, only 3.07% (95%CI: 2.71, 3.48) developed CRC. For woman the cumulative incidence for CRC at age 80 was 9.54% (95%CI:7.64, 12.11), 6.57% (95%CI:5.80, 7.33), 4.30% (95%CI:3.83, 4.74), 2.55% (95%CI:2.21, 2.92) for individuals in the ≥99th, 80th-99th, 20th-80th, and 0-20th percentile, respectively. The PRS groupings showed similar behavior across both ancestry groups (see Supplementary Figure 2 and Supplementary Figure 3).

At age 50, when the national screening program begins in Denmark, the cumulative incidence is 0.20% for men and 0.21% for women^36^. In the 20-80^th^ percentile, the same incidence is reached also at age 50. By contrast, individuals in the ≥99th percentile reach this incidence earlier, at ages 45 for males and 44 for females, and those in the 80^th^-99th percentile group reach it at age 47. In contrast, for the lowest-risk group (0-20th percentile), the same incidence is not reached until age 54. A similar pattern was observed between ages 45 and 60, where individuals with a high PRS reached the incidence levels of low-PRS individuals roughly 10 years earlier (see Supplementary Table 2).

Cox models showed that compared to individuals in the 20-80^th^ percentile, individuals in the 80-99^th^ and ≥99^th^ percentiles had significantly higher risk of developing CRC, with hazard ratio (HR) of 1.55 (95%CI: 1.48-1.62, p<0.001) and 2.22 (95%CI: 1.92-2.57, p<0.001), respectively. In contrast, individuals in the 0-20^th^ percentile had a reduced risk (HR: 0.55, 95%CI: 0.51-0.59, p<0.001). Males had a higher risk of developing CRC compared to females with a HR of 1.20 (95%CI: 1.15-1.25, p<0.001). There was no significant interaction between sex and PRS. There was no difference between European and non-European in the ancestry-stratified analysis (Supplementary Figure 4).

### Lifetime risk of adenoma in PRS groups and across sex

Cox models showed that a CRC PRS can stratify risk of adenomas almost as well as for CRC, see Supplementary Figure 4. Compared to individuals in the 20-80^th^ percentile, the risk of adenoma was higher for individuals in the 80-99^th^ and ≥99^th^ percentiles with HR of 1.44 (95%CI: 1.37-1.51, p<0.001) and 1.92 (95%CI: 1.64-2.25, p<0.001), respectively, and lower for individuals in the 0-20^th^ percentile (HR: 0.70, 95%CI: 0.66-0.74, P=p<0.001). Males had a higher risk of developing adenomas compared to females with a HR of 1.43 (95%CI: 1.37-1.49, p<0.001). There was no significant interaction between sex and PRS.

### Association of PRS with lifetime CRC risk across tumor characteristics

Cause-specific Cox models showed that, across histology types and CRC sites, individuals in the ≥99th and 80-99th percentiles had higher CRC risk compared to the reference group, while those in the 0-20th percentile had lower risk, see Figure 3. Exceptions were observed in small subgroups, such as unspecified colon site CRC (N=10), where sample size was low.

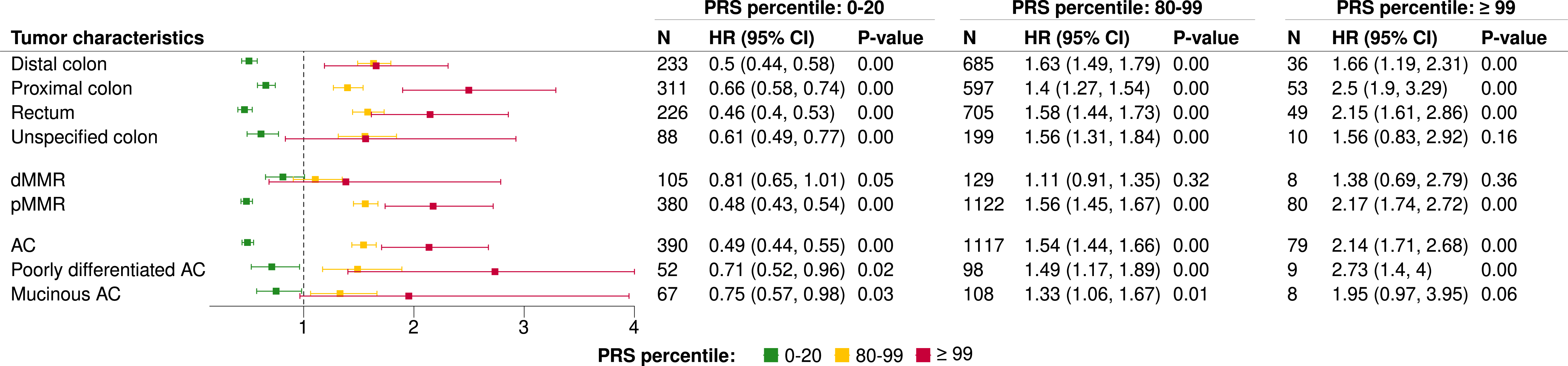

PRS also stratified risk among individuals with pMMR tumors but not among the 623 individuals with dMMR tumors. For dMMR tumors, no differences were observed between high-percentile groups and the reference group. Individuals in the 0-20th percentile had lower risk (HR=0.81, 95% CI=0.65-1.01, p=0.05), but still markedly higher than pMMR tumors in the same percentile (HR=0.48, 95% CI=0.43-0.54, p<0.001).

Cumulative incidence analyses confirmed that PRS stratifies risk in pMMR but not in dMMR tumors (Supplementary Figure 5). Demographics, clinical, and tumor characteristics for individuals with pMMR tumors and dMMR tumors are provided in Supplementary Table 3.

### CRC and adenoma risk prediction

Screening invitations were sent to 36,478 individuals in the CHB cohort, of whom 19,416 participated at least once. A total of 35,145 screenings (FIT tests) were conducted meaning that they on average had participated 1.81 times. Among the participants, 996 individuals were diagnosed with CRC and 1,715 with adenoma, with 296 and 1,396 of these cases identified through screening (positive FIT followed by positive colonoscopy), respectively. Using the screening participants three analyses were conducted. Summary statistics for all analyses are reported in Supplementary Table 4 and boxplots of fold-wise performance are presented in Figure 4.

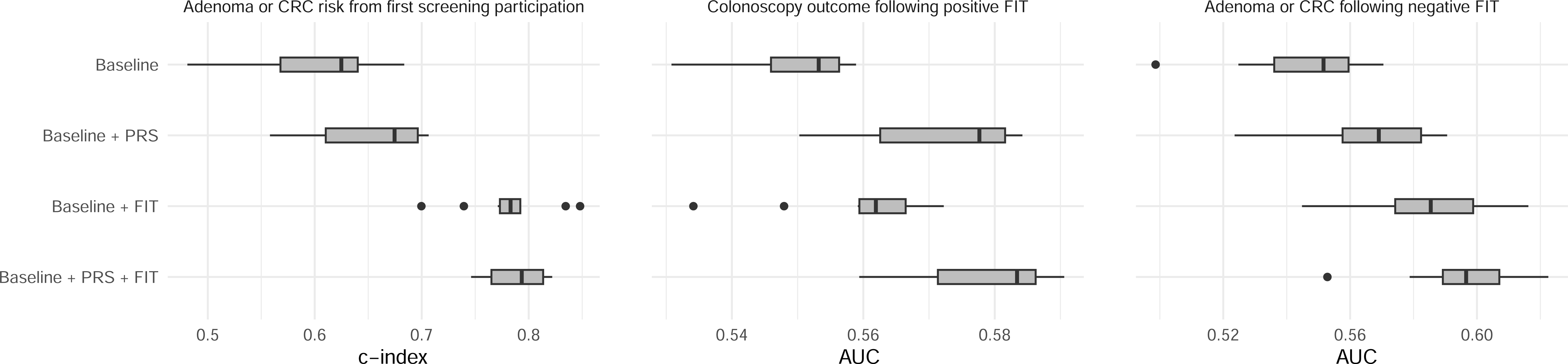

First, risk of adenoma or CRC was predicted from the time of first screening. A total of 3,547 participants were aged ≤56 years at screening initiation, of whom 281 were diagnosed with adenoma or CRC by the end of follow-up. Compared to the baseline + FIT model (median C-index=0.78, IQR=0.02), adding PRS (median C-index=0.79, IQR=0.05) did not markedly improve predictive performance.

Second, colonoscopy outcomes (1,660 positive vs. 1,625 negative) were predicted among FIT-positive participants. Predictive performance for colonoscopy outcomes was generally low (median AUC ≤ 0.59). Overall, the PRS + baseline model (median AUC=0.58, IQR=0.02) showed slightly better performance than the FIT + baseline model (median AUC=0.56, IQR=0.01). Adding FIT to the PRS + baseline model (median AUC=0.58, IQR=0.02) only marginally increased the AUC.

In the third analysis, the prediction of adenoma or CRC (interval cancer) within two years following a negative FIT result was examined. Among the 15,561 participants with only negative FIT results, 362 individuals developed an adenoma or interval cancer. Overall model performance was modest (median AUC<=0.60). While the FIT + baseline model (median AUC = 0.59, IQR=0.03) performed slightly better than the PRS + baseline model (median AUC=0.57, IQR=0.03), adding PRS to the FIT + baseline model resulted in a modest improvement in predictive performance (median AUC=0.60, IQR=0.02).

## Discussion

In this study, we estimated CRC PRS for 112,204 individuals in the CHB cohort and our findings highlight both the potential and the limitations of PRS in CRC screening. PRS effectively stratified lifetime risk of adenomas and CRC independent of sex and ancestry, identifying high-risk individuals who reached average population CRC risk around a decade earlier compared to those with low risk. However, its lack of association with dMMR tumors indicates important biological heterogeneity not captured by the current PRS. Moreover, PRS did not improve predictive performance when added to FIT at the first screening, and it showed only limited improvement in predicting negative colonoscopy outcomes among FIT-positive participants or the occurrence of adenoma or CRC within two years after a negative FIT.

Our study showed that PRS stratifies lifetime risk of adenomas and CRC, with individuals in the highest percentile reaching risks four to five times greater than those in the lowest. These findings are consistent with the only other study that recalibrated lifetime CRC risk to the general population^23^. Their PRS achieved stronger stratification, likely because they restricted analyses to European ancestry and applied methods incorporating linkage disequilibrium. In contrast, we used a simpler weighted-sum approach based on 205 variants, which is known to be outperformed by methods incorporating linkage disequilibrium ^16^. However, the method allows inclusion of both European and non-European ancestry groups, ensuring representativeness for the Danish population and a more real picture of what is possible now.

An important consideration before moving towards clinical validation and ultimately implementation is whether PRS can stratify risk across all individuals, independent of tumor characteristics. We found that PRS identified high and low risk individuals across CRC sites and histology, but not among dMMR tumors. About 10-15% of CRCs are dMMR, caused by either sporadic events or inherited variants (e.g., HNPCC). Our findings align with one study showing that PRS was predictive in the general population but not in HNPCC ^25^, suggesting it also applies to sporadic cases. MMR genes repair replication errors, and pathogenic variants interrupt this process, causing somatic variants to accumulate and increasing CRC risk. Such MMR mechanisms might be less influenced by the cumulative effect of common low-risk variants, which PRS captures. In contrast, most CRCs are pMMR, where polygenic factors likely play a larger role. Since individuals with HNPCC are already offered more regular screening, the main challenge lies with sporadic dMMR tumors or undiagnosed HNPCC cases, which constitute the majority of dMMR tumors.

These findings raise the question of whether PRS could inform the starting age or intensity of CRC screening. The appropriate age to begin CRC screening remains debated and varies across countries^4^. Our results show that cumulative incidence differs between PRS groups already at early ages, supporting differentiated starting ages based on genetic risk, in line with prior studies^17,18,22^.

Screening intensity is also under discussion. For example, an ongoing breast cancer trial screens high-risk individuals more frequently^40^. The differences in cumulative incidence across PRS groups suggest that PRS could be used to stratify individuals into high- and low-risk groups. However, further studies are needed to determine the optimal thresholds for PRS groupings, as there is a linear association between PRS and lifetime CRC risk without natural cutoffs. These studies should balance clinical benefits with potential harms and costs to establish evidence-based, PRS-guided screening strategies.

This study, the largest to date examining the utility of PRS for risk-based CRC screening, provides realistic lifetime risk estimates for CRC and adenomas. It is ancestry-inclusive and population-recalibrated, incorporating tumor features and the previously unexplored tumor MMR status. However, the study uses a hospital cohort, leading to a higher proportion of CRC cases compared to the general population, this was addressed by recalibrating to the general Danish population. Due to the lack of available incidence and prevalence estimates for some variables (e.g., adenomas), recalibration of cumulative incidences for these variables was not possible. Another challenge with this cohort, as with other biobanks, is the exclusion of individuals who died before its establishment (CHB, 2009), which could introduce selection bias. However, a simulation study shows this is negligible if post-disease fatality is independent of the exposure^41^. Since the PRS affects disease risk but likely not post-disease mortality, this limitation is unlikely to impact our estimates. Finally, predictive models showed only limited improvement when adding PRS to FIT. However, this analysis was limited by short follow-up (the program was initiated in 2014) and a relatively young cohort with few CRC and adenoma cases. Longer follow-up, larger sample sizes, and models allowing more complex interactions, including dynamic risk prediction, should be explored to assess the utility of PRS in predicting adenoma and CRC risk.

In conclusion, PRS effectively stratifies lifetime CRC and adenoma risk, except for dMMR tumors, and shows promise for guiding risk-based screening initiation and intensity. When combined with FIT in predictive modeling, it provided only limited additional improvement in identifying false positives and false negatives.

## Data availability

Data from CHB is available on reasonable request. The risk variants and their weights used to estimate the PRS are from a publicly available GWAS and can be downloaded from the PGS Catalog^42^ at https://www.pgscatalog.org/score/PGS003850/.

## Acknowledgments

We acknowledge all CHB participants and the clinicians in the gastroenterology, surgical, and oncology departments across the Capital Region who cared for patients whose residual blood samples contributed to the collection of CRC samples for CHB. We also thank the staff at the Clinical Immunology departments across the Capital Region and the Biobank Unit at Copenhagen University Hospital for their contributions to the establishment and maintenance of the CHB. We furthermore thank deCODE genetics for performing the genotyping and imputation of the biological samples.

This work was supported by the Danish Cancer Society (R374-A2269) and the Aalborg University Excellence Program to Anne Krogh Nøhr.

CHB is supported by the Department of Clinical Immunology, Rigshospitalet, Copenhagen University Hospital, Copenhagen, Denmark and by grants from Novo Nordisk Foundation (NNF23OC0082015, NNF17OC0027594) and Rigshospitalet Research Council (Framework grant).

The Danish Blood Donor Study (DBDS) is funded by an annual grant from Bio- and Genome Bank Denmark. The initiation of DBDS was supported by the Danish Administrative Regions (02/2611) and the Danish Council for Independent Research (09–069412). Additionally, the DBDS is funded by the Novo Nordisk Foundation (NNF23OC0082015, NNF17OC0027864, and NNF17OC0027594).

## Consortia

### DBDS Genomic Consortium

Karina Banasik, Jakob Bay, Andrea Barghetti, Mette Skou Bendtsen, Jens Kjærgaard Boldsen, Søren Brunak, Nanna Brøns, Alfonso Buil Demur, Johan Skov Bundgaard, Lea Arregui Nordahl Christoffersen, Maria Didriksen, Khoa Manh Dinh, Joseph Dowsett, Christian Erikstrup, Josephine Gladov, Daniel Gudbjartsson, Thomas Folkmann Hansen, Dorte Helenius Mikkelsen, Lotte Hindhede, Henrik Hjalgrim, Jakob Hjorth von Stemann, Bitten Aagaard Jensen, Ingileif Jónsdóttir, Kathrine Kaspersen, Bertram Dalskov Kjerulff, Lisette Kogelman, Mette Kongstad, Susan Mikkelsen, Christina Mikkelsen, Line Hjorth Sjernholm Nielsen, Janna Nissen, Mette Nyegaard, Sisse Rye Ostrowski, Frederikke Byron Pedersen, Ole Birger Pedersen, Liam James Elgaard Quinn, Þórunn Rafnar, Palle Duun Rohde, Klaus Rostgaard, Andrew Joseph Schork, Michael Schwinn, Erik Sørensen, Kari Stefansson, Hreinn Stefánsson, Jacob Træholt, Unnur Þorsteinsdóttir, Mie Topholm Bruun, Henrik Ullum, Thomas Werge & David Westergaard.

### DCB Research Consortium

Michael Patrick Achiam, Christian Brieghel, Rasmus Froberg Brøndum, Nanna Brøns, Martin Bøgsted, Olafur B. Davidsson, Jojo Biel-Nielsen Dietz, Karen Dybkær, Christian Erikstrup, Kirsten Grønbæk, Mathias Ærendahl Heldbo, Hannes Helgason, Jens G Hillingsø, Lotte Hindhede, Henrik Hjalgrim, Simon Husby, Estrid Høgdall, Tereza Fait Kadlec, Bertram Kjerulff, Hildur Knutsdottir, Ragnar Pétur Kristjánsson, Ulrik Lassen, Cecilie Velsoe Maeng, Magnus K Magnusson, Pei Meng, Carsten Utoft Niemann, Anne Krogh Nøhr, Sisse R Ostrowski, Inge Søkilde Pedersen, Ole Birger Pedersen, Hans-Christian Pommergaard, Liam Quinn, Andreas Røder, Michael Schwinn, Simon N Stacey, Hein V Stroomberg, Erik Sørensen, Rebecca Svanberg Teglgaard, Steffen UIlltz Thorsen, Unnur Thorsteinsdottir & Bragi G Walters.

