## Supplementary Material for "Combining genome-wide polygenic scores with registry data for colorectal cancer risk-based screening"

#### Supplementary Material 1

EDTA whole blood samples were genotyped by deCODE Genetics using Illumina's Infinium Global Screening Array v1 or v3. Genotype data underwent standard quality control, including removal of duplicate samples, samples failing sex checks, and samples with missingness >5%. Variants with missingness >10% or a Hardy-Weinberg equilibrium exact test p-value < 0.00001 were removed. The QCed genotype data were imputed using deCODE genetics in-house imputation work-flow[35] and an in-house reference panel of approximately 50,000 individuals of mixed ancestry among these around 10,800 Danish individuals.

Principal components (PCs) were estimated from the autosomal, unimputed genotype data. A random subset comprising 30% of unrelated individuals (defined as KING kinship coefficient < 0.044) was selected. Basic quality control was applied to this subset, excluding variants with >1% missingness, Hardy-Weinberg equilibrium p-values < 0.0001, and minor allele frequency <1%. Singular value decomposition (SVD) was used iteratively (up to five iterations) to remove individuals most dissimilar to the rest. PCs were then estimated on the remaining individuals using PLINK 2.0. Subsequently, all remaining individuals were projected into the principal component space defined by the unrelated subset.

Ancestry was estimated using principal component analysis (PCA) on unrelated individuals, with related individuals and 1000 Genomes Project (1KG) samples projected into the same space. To enhance clustering, Uniform Manifold Approximation and Projection (UMAP) was applied to the PCA results. Individuals genetically similar to 1KG Europeans were assigned to the “EUR” cluster using Mahalanobis distance (5 standard deviations (SD)). The same approach was applied to other major ancestry groups (African, South Asian, East Asian, and Admixed American), with genetic similarity defined as within three SDs of the Mahalanobis distance to the respective 1KG reference panel. Individuals not genetically aligned with any 1KG ancestry cluster were categorized as admixed. Finally, individuals with both parents born in Greenland who did not cluster with EUR were assigned Greenlandic (GRL) ancestry.

The first two PCs, colored by genetic ancestry, are shown in Figure 1, and the variance explained by the first nine PCs is visualized in Figure 2.

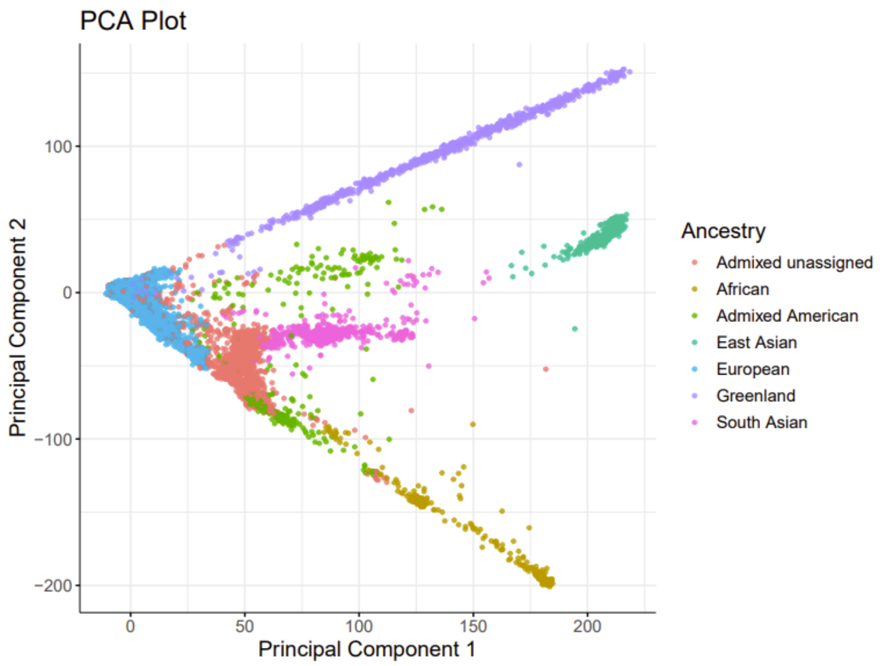


**Figure 1:** PC1 and PC2 colored by genetic ancestry

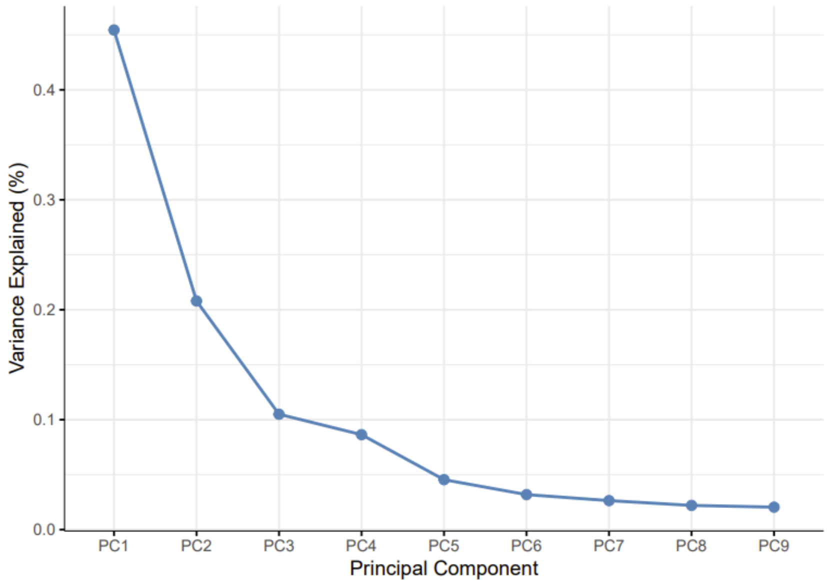


**Figure 2:** Variance explained by the first nine PCs

#### Supplementary Material 2

**Method**

Linear and non-linear association between PRS and lifetime risk of CRC were evaluated for Cox proportional hazards models. Thus, follow-up began at the entry date 1978-10-01 (In 1998 ICD-O coding was introduced in CAR and DNPR achieved complete nationwide coverage) or at birth for individuals born after that date. For cases, follow-up ended at age of first CRC diagnosis. For controls, follow-up ended at age of death from a cause other than CRC, age at last record in the registries or age 80. All models included left truncation at a CRC-relevant entry date (listed in Supplementary Table 1) to avoid immortal time bias. Each model was adjusted for the first four principal components, birth year, and chip version. First a linear model was made. Secondly, cubic splines models were evaluated with 3-7 knots, this is an example of a 3-knot model. For all models Akaike Information Criterion (AIC) was estimated, asymptotic likelihood ratio test (LRT) with a chi-square distribution, and visual inspection was used to compare the linear and the spline model.

**Results**

Linear and spline-based Cox models (with 3 to 7 knots) were fitted to the data to examine if there were any natural cutoffs for PRS group definitions in the associations between PRS and lifetime risk of CRC. Estimation of each models AIC showed that the spline model with 3 knots had the lowest AIC (197195.1), see Table below. This was supported by the LRT, which indicated that the 3-knot spline model was significantly better than the linear model (p = 0.0001), while models with more than three knots did not provide a significantly better fit. However, visual inspection of the 3-knot spline model compared to the linear model revealed only minor differences in the fitted curves (see Supplementary Figure 1). Thus, there were no clear clinical advantages to choosing the more complex spline model over the linear model. Furthermore, the wider CIs at the extremes of the PRS distribution reflect the limited number of individuals with very low or very high PRS, raising further questions about the robustness of the spline model in these regions. Consequently, the linear model was selected for subsequent analyses. As no natural cutoffs for PRS groupings emerged from the model, individuals were categorized into PRS groups as follows: <1%, 1-20%, 20-80% (reference), 80-99% or ≥99%. These groupings were aligned with those used in Tamlander et al. 2024[23] to enable direct comparison.

| Model | Knots | AIC | LRT |
|---|---|---|---|
| **Linear** | - | 197208.0 | |
| **Spline** | **3** | **197195.1** | **0.0001** |
| | 4 | 197197.1 | 0.75 |
| | 5 | 197196.9 | 0.15 |
| | 6 | 197199.1 | 0.64 |
| | 7 | 197200.4 | 0.40 |

#### Supplementary Material 3

A subset of individuals was linked to DDSCE, and three analyses were conducted to assess whether the addition of PRS to FIT, compared with FIT alone, enhanced the performance of:

1. Prediction of adenoma or CRC risk from the time of an individual's first screening participation.
2. Prediction of colonoscopy outcomes (positive vs. negative findings) among participants with a positive FIT.
3. Prediction of adenoma or CRC within two years following a negative FIT result.

In all three analyses model performance was evaluated for four distinct predictor sets: (1) a baseline set including age, sex, principal components 1-4 (PC1-PC4), and genotyping chip; (2) baseline plus PRS; (3) baseline plus FIT value; and (4) baseline combined with both PRS and FIT. In all analysis FIT was included as a continuous variable and linear prediction models were chosen due to the limited sample sizes.

**Analysis 1** assessed CRC or adenoma risk prediction using Cox proportional hazards models. The cohort included participants aged 56 years or younger at the time of their first FIT test. Time-to-event modeling was conducted using the coxph() function from the survival package. Follow-up time began at the date of the first FIT and ended at the earliest of CRC/adenoma diagnosis, death, or censoring date. 10-fold cross-validation was performed using the survcompare package, and predictive accuracy was evaluated using c-index.

For **Analyses 2 and 3**, logistic regression models were fitted using the glm() function from base R. 10-fold cross-validation was performed using the caret package, and predictive accuracy was assessed using the AUC computed with the pROC package.

**Analysis 2** focused on participants with a positive FIT result and a follow-up colonoscopy. If a participant had more than one positive FIT result the first was used. Cases were defined as participants with a positive colonoscopy, while controls included those with a negative colonoscopy result. Participants without colonoscopy data were excluded.

**Analysis 3** included participants with negative FIT results and no prior diagnosis of adenoma or CRC. Cases were defined as individuals diagnosed with adenoma or CRC within two years following a negative FIT result. Controls were selected from a pool of eligible participants who (i) had a negative FIT and no prior diagnosis of CRC or adenoma, and (ii) were not diagnosed with adenoma, CRC or died within the two years following the negative FIT. Three controls were matched to each case from this pool based on sex and age at the time of the FIT test. For cases, the FIT administered closest to the time of diagnosis was selected. For controls with multiple FITs, the FIT taken at the age corresponding to the matched case was used. Participants with an adenoma or CRC diagnosis prior to all FIT tests were excluded.

### Supplementary Table 1

| Outcome | Analysis | Left truncation date | Argument |
|---|---|---|---|
| **CRC** | Cox proportional hazards models and Cumulative incidence models | 1978-01-01 | CAR was established in 1942, but data quality improved in 1978 because ICD-O coding was introduced. |
| **Adenoma** | Cox proportional hazards models and Cumulative incidence models | 1978-01-01 | DNPR achieved complete nationwide coverage in 1978. |
| **CRC histology** | Cause-specific Cox proportional hazards models | 2009-10-01 | Data on histology have been available in DCCG since this date. |
| **CRC MMR status** | Cause-specific Cox proportional hazards models | 2009-10-01 | Data on MMR protein expression have been available in DCCG since this date. |
| **CRC site** | Cause-specific Cox proportional hazards models | 2001-07-01 | Data on tumor site have been available in DCCG since this date |

#### Supplementary Table 2

| Age | Female | | Male | |
|---|---|---|---|---|
| | Population incidence CRC[1] | Age (years) at incidence in this cohort | Population incidence CRC[1] | Age (years) at incidence in this cohort |
| **45 years** | 0.10% | | 0.09% | |
| PRS≥ 99% | | 39 | | 39 |
| PRS 80–99% | | 42 | | 42 |
| PRS 20–80% | | 45 | | 45 |
| PRS 0–20% | | 48 | | 49 |
| **50 years** | 0.21% | | 0.20% | |
| PRS≥ 99% | | 44 | | 45 |
| PRS 80–99% | | 47 | | 47 |
| PRS 20–80% | | 50 | | 50 |
| PRS 0–20% | | 54 | | 54 |
| **55 years** | 0.41% | | 0.43% | |
| PRS≥ 99% | | 49 | | 50 |
| PRS 80–99% | | 52 | | 52 |
| PRS 20–80% | | 55 | | 55 |
| PRS 0–20% | | 59 | | 59 |
| **60 years** | 0.79% | | 0.87% | |
| PRS≥ 99% | | 53 | | 54 |
| PRS 80–99% | | 56 | | 56 |
| PRS 20–80% | | 60 | | 60 |
| PRS 0–20% | | 65 | | 65 |

[1] Incidence estimates for Denmark from Global Burden of Disease (GBD) 2021 data.

### Supplementary Table 3

| Characteristic | MMR deficient N = 623 | MMR proficient N = 3,895 |
|---|---|---|
| **Diagnosis group** | | |
| Colon (C18,26) | 603 (97%) | 2,586 (66%) |
| Rectum (C19-20) | 20 (3%) | 1,309 (34%) |
| **Gender** | | |
| Female | 408 (65%) | 1,493 (38%) |
| Male | 215 (35%) | 2,402 (62%) |
| **Birth year** | 1941 (1936, 1946) | 1944 (1937, 1951) |
| **Year of diagnosis** | 2016 (2013, 2018) | 2015 (2013, 2017) |
| **CRC site** | | |
| Distal colon | 63 (14%) | 1,318 (38%) |
| Proximal colon | 303 (68%) | 575 (17%) |
| Rectum | 20 (4.5%) | 1,309 (38%) |
| Unspecified colon | 61 (14%) | 236 (7%) |
| Unknown | 176 | 457 |
| **ECOG Performance Status[2]** | | |
| 0 | 179 (42%) | 1,372 (56%) |
| 1 | 151 (35%) | 686 (28%) |
| 2 | 71 (17%) | 270 (11%) |
| +3 | 29 (7%) | 118 (5%) |
| Unknown | 193 | 1,449 |
| **Histology** | | |
| Adenocarcinoma | 301 (48%) | 3,339 (86%) |
| Unknown or carcinoma (Medullary, Signet ring cell, Undifferentiated) | 67 (11%) | 37 (1%) |
| Poorly differentiated adenocarcinoma | 138 (22%) | 212 (5%) |
| Mucinous adenocarcinoma | 117 (19%) | 307 (8%) |
| **Adenoma diagnosis** | | |
| Adenoma | 119 (19%) | 762 (20%) |
| No adenoma | 504 (81%) | 3,133 (80%) |
| **Ancestry** | | |
| European | 576 (92%) | 3,600 (92%) |
| Non-European | 47 (8%) | 295 (8%) |
| **PRS strata** | | |
| 0-20 | 105 (17%) | 380 (10%) |
| 20-80 | 381 (61%) | 2,312 (59%) |
| 80-99 | 129 (21%) | 1,123 (29%) |
| ≥99 | 8 (1%) | 80 (2%) |

[1] n (%); Median (Q1, Q3), [2] ECOG = Eastern Cooperative Oncology Group

#### Supplementary Table 4

| Analysis 1: Risk of adenoma or CRC from the time of first screening | | |
|---|---|---|
| **Model** | **Median C-index** | **IQR** |
| Baseline | 0.625 | 0.072 |
| Baseline + PRS | 0.675 | 0.086 |
| Baseline + FIT | 0.783 | 0.019 |
| Baseline + PRS + FIT | 0.794 | 0.048 |
| **Analysis 2: Colonoscopy outcome among FIT-positive participants** | | |
| **Model** | **Median AUC** | **IQR** |
| Baseline | 0.553 | 0.010 |
| Baseline + PRS | 0.578 | 0.019 |
| Baseline + FIT | 0.562 | 0.007 |
| Baseline + PRS + FIT | 0.583 | 0.015 |
| **Analysis 3 - Adenoma or CRC within two years following a negative FIT** | | |
| **Model** | **Median PR AUC** | **IQR** |
| Baseline | 0.028 | 0.001 |
| Baseline + PRS | 0.031 | 0.003 |
| Baseline + FIT | 0.032 | 0.005 |
| Baseline + PRS + FIT | 0.034 | 0.003 |

#### Supplementary Figure 1

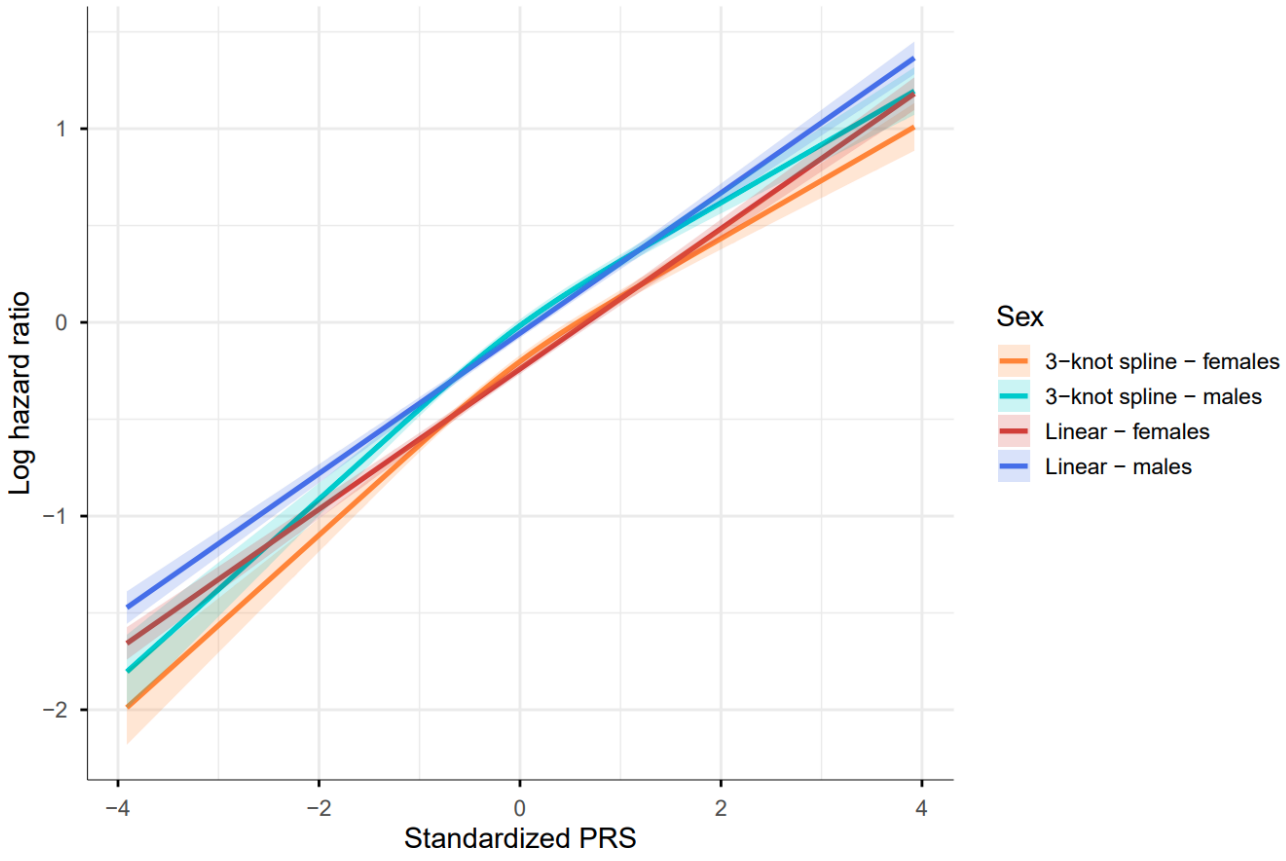


Log hazard ratios (HR) with 95% CIs as a function of standardized PRS, estimated using both linear and spline-based (3-knot) Cox proportional hazards models for men and women.

#### Supplementary Figure 2

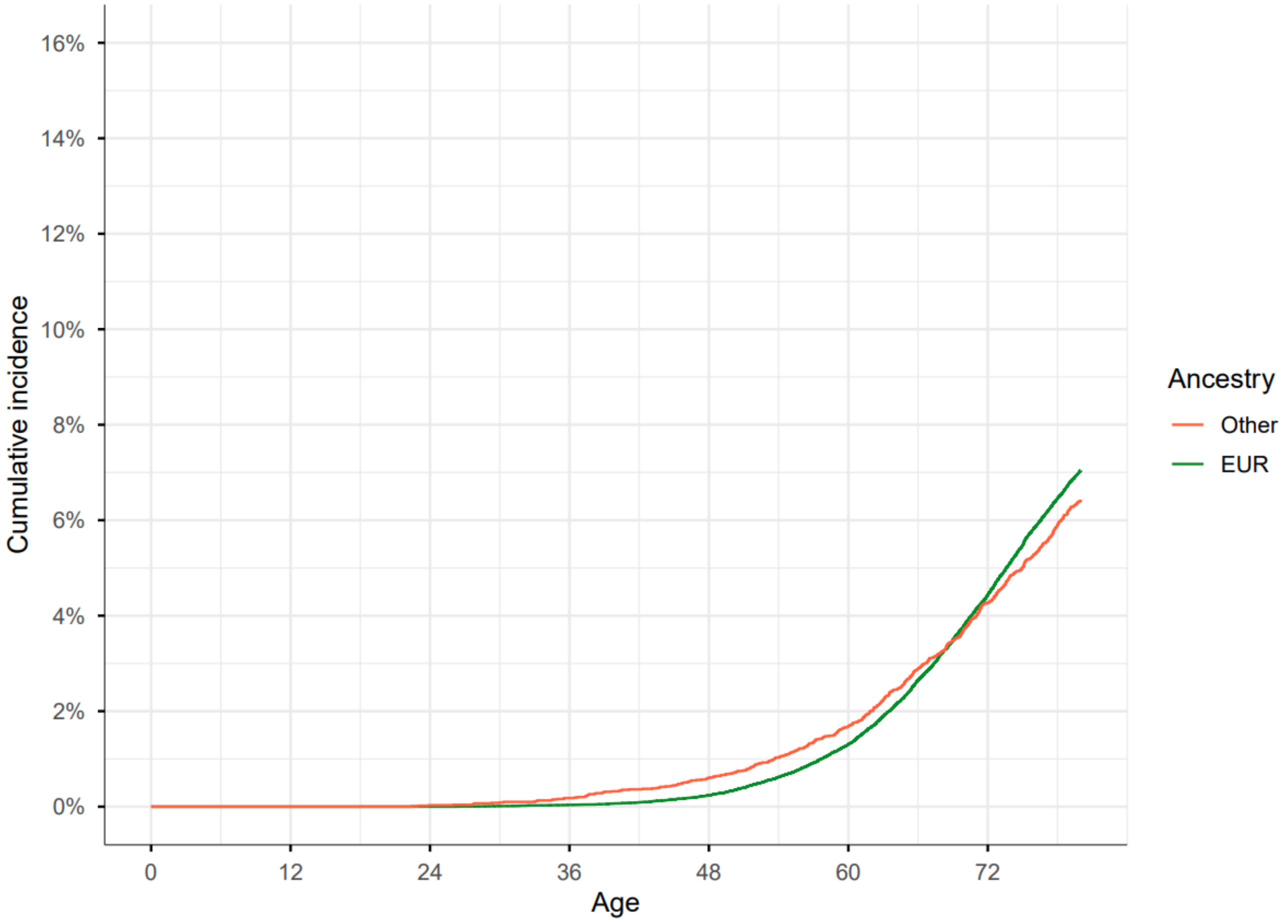


Cumulative incidence of lifetime colorectal cancer risk, stratified by European and non-European ancestry. Note: The cumulative incidence has not been recalibrated to the Danish population, as this would require the assumption that incidence, prevalence, and mortality estimates are directly comparable across the two groupings, which was not deemed appropriate.

#### Supplementary Figure 3

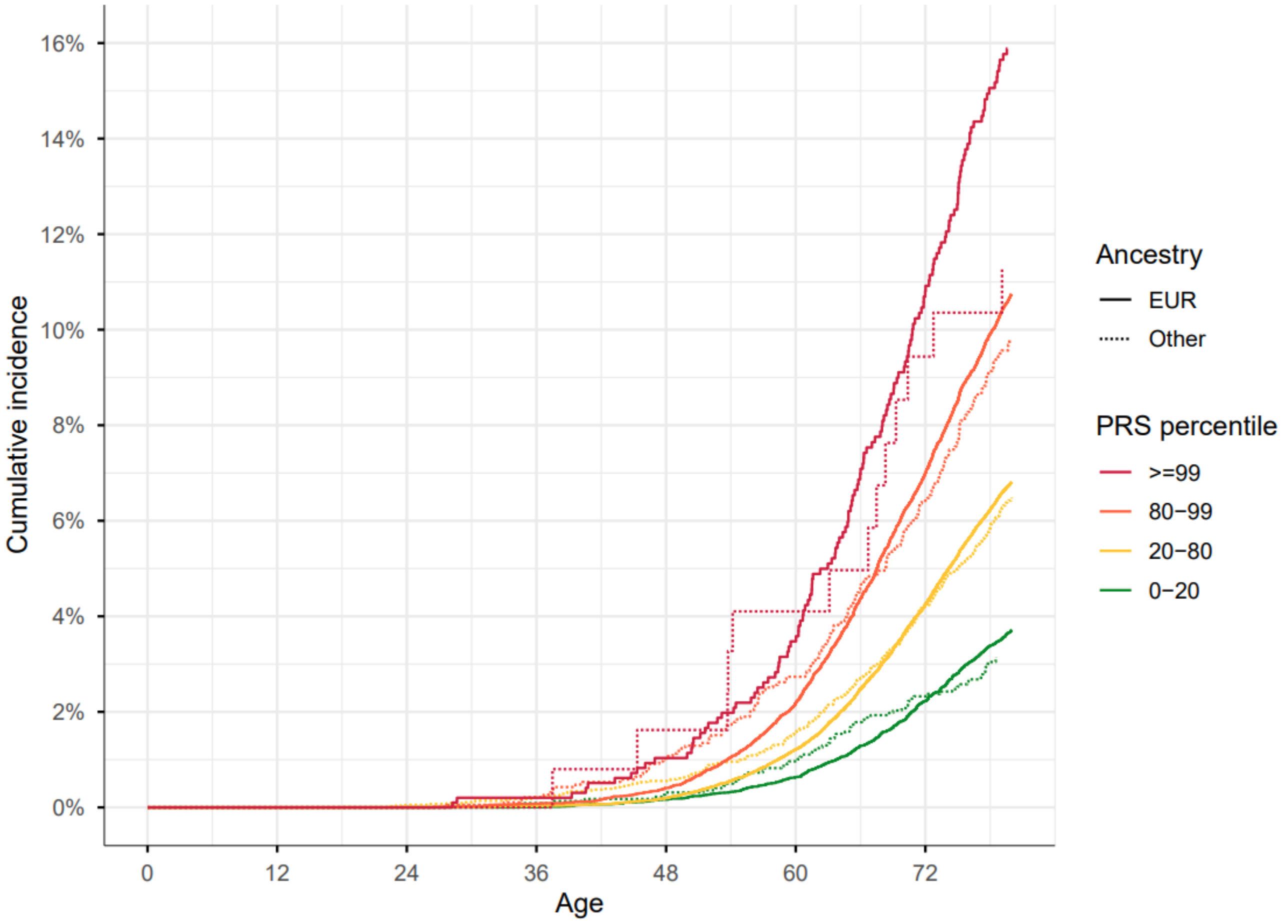


Cumulative incidence of lifetime colorectal cancer risk across PRS groups, stratified by European and non-European ancestry.

#### Supplementary Figure 4

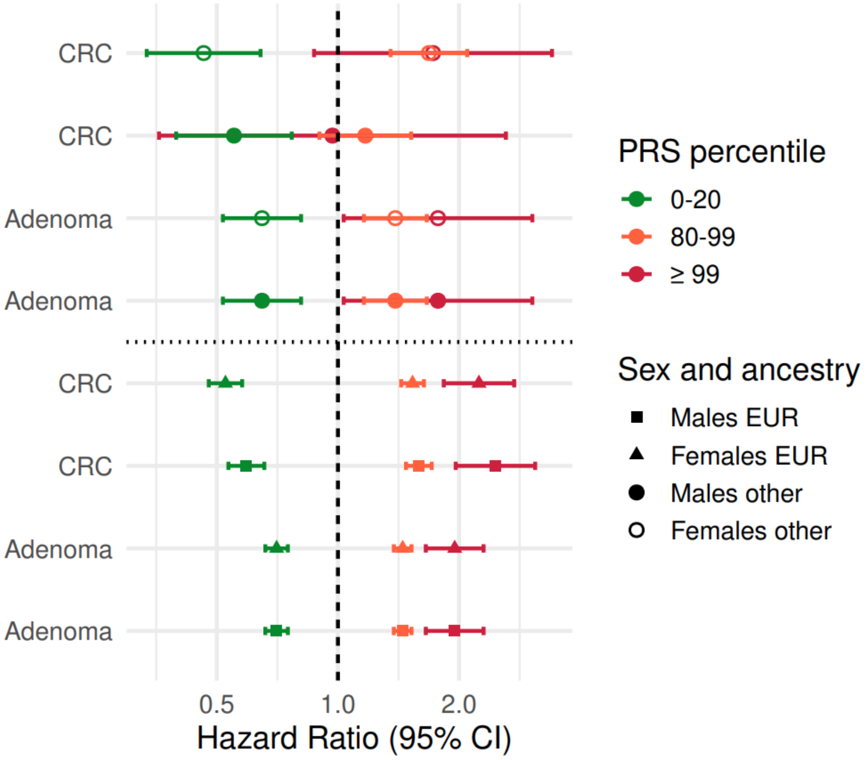


HR with 95% CIs for lifetime risk of colorectal cancer and adenoma across PRS groupings, stratified by sex.

#### Supplementary Figure 5

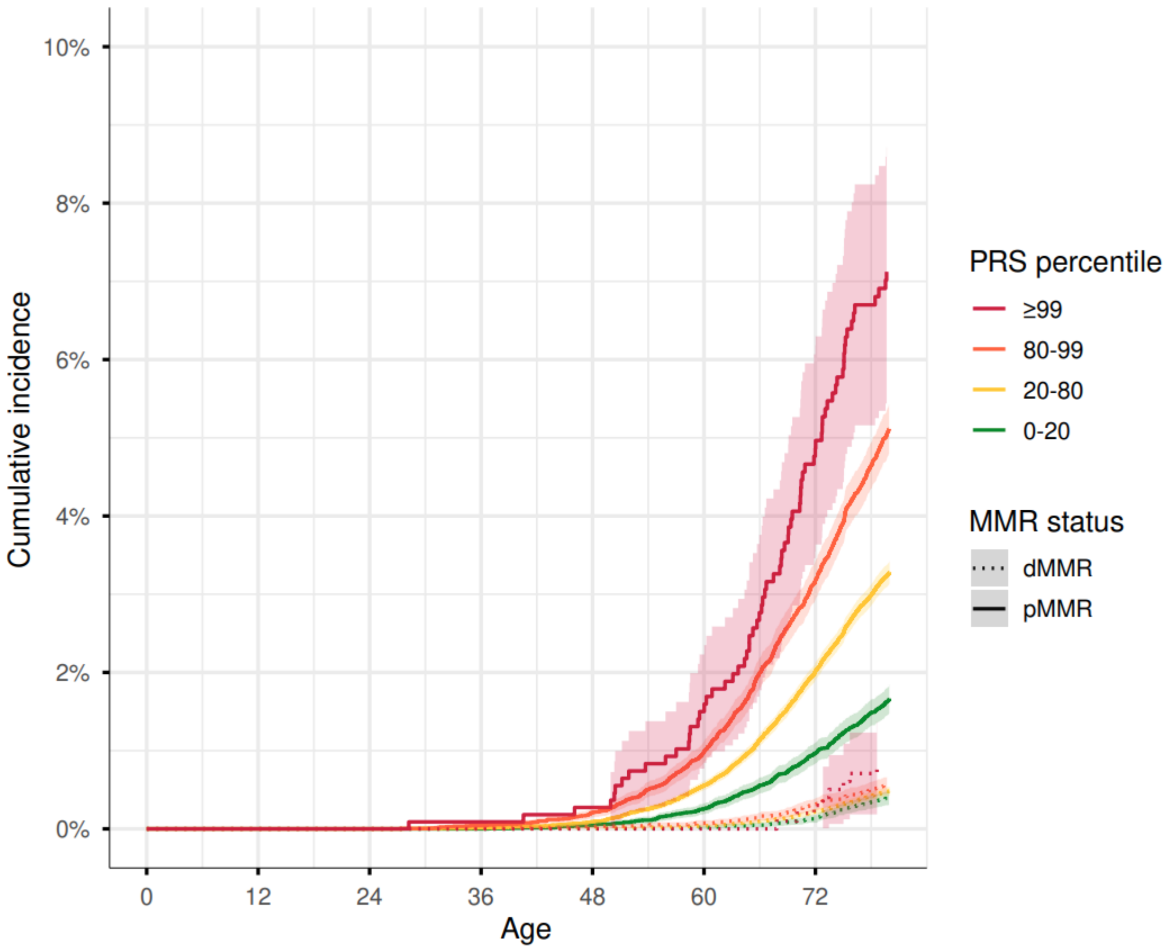


Cumulative incidence with 95% CIs of lifetime colorectal cancer risk across PRS groupings, stratified by MMR status. Note: The cumulative incidence has not been recalibrated to the Danish population, as this would require the assumption that incidence, prevalence, and mortality estimates are directly comparable across the two groupings, which was not deemed appropriate.
